# Risk of Clonal Hematopoiesis of Indeterminate Potential after Cancer Radiation Therapy

**DOI:** 10.1101/2024.09.27.24314321

**Authors:** Shelby A. Crants, Sydney S. Olson, Yajing Li, Cosmin A. Bejan, Caitlyn Vlasschaert, Taralynn M. Mack, Yash Pershad, Ashwin Kishtagari, Sarah C. Reed, Sarah Croessmann, Dan M. Roden, Travis J. Osterman, Eric T. Shinohara, Alexander G. Bick, Ben H. Park, Leo Y. Luo

**Affiliations:** Vanderbilt University School of Medicine, Nashville, Tennessee, USA; Division of Genetic Medicine, Department of Medicine, Vanderbilt University Medical Center, Nashville, Tennessee, USA; Department of Biostatistics, Vanderbilt University Medical Center, Nashville, Tennessee, USA; Department of Biomedical Informatics, Vanderbilt University Medical Center, Nashville, Tennessee, USA; Department of Medicine, Queen’s University, Kingston, Ontario, Canada; Division of Hematology and Oncology, Department of Medicine, Vanderbilt University Medical Center, Nashville, Tennessee, USA; Division of Clinical Pharmacology, Department of Medicine, Vanderbilt University Medical Center, Nashville, Tennessee, USA; Department of Radiation Oncology, Vanderbilt University Medical Center, Nashville, Tennessee, USA

## Abstract

Clonal hematopoiesis of indeterminate potential (CHIP) is a condition associated with aging and increased risk of hematologic malignancies and chronic diseases. While radiation therapy (RT) has been implicated as a risk factor for CHIP, the specific radiation parameters influencing the development of CHIP remain unclear. This study aimed to investigate the association between RT and CHIP and identify relevant radiation parameters affecting CHIP risk. We conducted a retrospective cohort study of cancer patients with RT exposure from an institutional biobank. DNA sequencing was performed to detect mutations in 22 CHIP-associated genes. Multivariable logistic regression models were used to evaluate the association between clinical characteristics, RT parameters, and CHIP. We identified 736 cancer patients with RT exposure and compared their risk of CHIP to a control cohort of 13,605 individuals. RT was found to be an independent risk factor for developing CHIP (OR 1.39, 95% CI 1.08-1.77, p=0.009). The prevalence of CHIP in RT patients was 22.8%. Compared to controls, RT patients showed significantly increased mutations in *DNMT3A*, *PPM1D*, *TP53,* and *BRCC3.* A positive correlation was observed between CHIP risk and biological equivalent dose of radiation. Stereotactic radiation was also associated with increased CHIP prevalence (OR 2.24, 95% CI 1.14-4.33, p=0.02). This study demonstrates that RT is associated with an increased risk of CHIP development, particularly mutations in DNA damage response genes. These findings have important implications for cancer care and long-term patient monitoring, emphasizing the need for further research into the mechanisms and consequences of RT-related CHIP.

**Key Points:**

- Radiation therapy significantly increases the risk of developing clonal hematopoiesis of indeterminate potential (CHIP) in cancer patients
- CHIP risk is higher in those who receive higher total dose and stereotactic radiation, with increased mutations in DNA damage repair genes

## Introduction

As modern cancer treatments improve patient survival, there is a growing recognition of the adverse effects on the normal organ functions from cancer therapies.^1^ Specifically, radiation therapy (RT) can cause various long-term side effects that are typically limited to the region of radiation exposure. For example, head and neck radiation can affect taste and swallowing, brain radiation may impact memory and cognition, and pelvic radiation can lead to bowel and bladder issues. A newly recognized sequela of RT is its effect on the genomic composition of the hematopoietic system. Recent studies have observed clonal expansion in hematopoietic stem cells (HSCs), a phenomenon termed clonal hematopoiesis, after cancer therapy such as RT and cytotoxic chemotherapy. ^2–6^

Within the spectrum of clonal hematopoiesis, clonal expansion of HSCs driven by somatic mutations in the absence of morphological evidence of a hematologic neoplasm is referred to as clonal hematopoiesis of indeterminate potential (CHIP).^7^ CHIP has been frequently observed in healthy older individuals.^8–10^ Although CHIP itself is not a malignant condition, clonal cells can serve as leukemic precursors, significantly increasing the risk of hematologic malignancies. ^11–13^ Patients with CHIP have a 0.5-1% risk per year of progressing to hematologic malignancy compared to individuals without CHIP.^7^ Furthermore, CHIP has been implicated in multiple chronic diseases associated with aging, such as cardiovascular diseases^14–17^, pulmonary diseases^18,19^, acute kidney injury^20^, chronic liver disease^21^, and other inflammatory conditions.^22,23^ While older age has been the dominant risk factor for developing CHIP, extrinsic factors resulting in DNA damage and replicative stress such as smoking, RT, and chemotherapy, have all been implicated as contributors to its pathogenesis. Various forms of radiation exposure, such as external beam radiation, radionuclide therapy, environmental radon, and human spaceflight, have all been associated with increased risk of CHIP.^2,6,24,25^

Although the association between RT and subsequent development of CHIP is known, it is largely unclear what RT parameters, such as radiation dose, radiation technique, and irradiated body site, may influence the process of CHIP development. This is, in part, due to limited granularity in the details of radiation data that can be found. Our large institutional DNA biobank linked with patient-level electronic health records (EHR) provided a unique opportunity to study the genotype-phenotype association between CHIP and RT exposure. The objectives of this study were to determine CHIP mutational landscape after RT, and to identify relevant radiation parameters associated with risk of CHIP.

## Methods

### Overview of Study Design

This protocol (IRB #220474) was approved by the Institutional Review Board (IRB) at Vanderbilt University Medical Center. This was a retrospective cohort study utilizing participants with blood samples in an institutional biobank (BioVU). ^26,27^ Patients who consented to having personal health information included in BioVU had DNA samples collected from blood remaining after routine clinical testing at the medical center. The blood samples are coupled to de-identified EHR data in the institution’s Synthetic Derivative (SD). The authors compared a cohort of RT-exposed patients from BioVU with a control cohort of BioVU patients who did not have exposure to RT.

### Cohort Identification

To identify the RT-exposed cohort, we interrogated the BioVU and SD database for patients billed for at least one Current Procedural Terminology (CPT) code associated with RT (see Supplemental Table 1) dated at least 6 months prior to the date of their blood draw to allow time for CHIP development. We further utilized natural language processing to identify patients who did not meet CPT code criteria but had at least one note in their EHR referencing “Prior radiation therapy:” that was not followed by “none” or “no” and was dated prior to the date of their BioVU sample blood draw to capture patients who received RT at other institutions. We manually reviewed the EHR to confirm that each identified patient had received RT and to extract demographic information, RT information, and site of malignancy where applicable. Patients were included if they were exposed to RT at least 6 months prior to the date of their blood draw (n = 736). The control cohort included 13,605 BioVU participants without RT exposure.^20,28^ Identical targeted DNA sequencing assay for CHIP mutations with error correction was performed on all individuals.

### DNA Sequencing and CHIP Identification

DNA from BioVU samples were sequenced using an institutional Clonal Hematopoiesis Sequencing Assay.^29^ The assay detects 22 frequently mutated CHIP genes that encompasses 95% of CHIP mutations in the TOPMed cohort.^9^ The genes include: *ASXL1, ASXL2, BRCC3, CBL, DNMT3A, ETNK1, GNAS, GNB1, IDH1, IDH2, JAK2, KIT, KRAS, MPL, NRAS, PPM1D, SETBP1, SF3B1, SRSF2, TET2, TP53,* and *U2AF1*. Putative somatic mutations were identified in the aligned sequencing reads using the Mutect2-GATK software package. The mutation filtering was applied to identify variants that have met at least 2% variant allele frequency (VAF) based on previously described criteria for CHIP identification.^30^ Variants with total read depth < 100, variant allele read depth < 3, and/or variant allele fraction below the 2% threshold for CHIP definition were removed.

### Statistical Analysis

Chi-squared analyses were performed to determine statistical significance of categorical variables, and p-values were derived from the chi-squared distribution. The association between CHIP and RT was evaluated using multivariable logistic regression models, adjusting for potential confounders of age, sex, race, and chemotherapy exposure, and p-values were calculated using two-tailed tests. Of note, smoking status was not included in the multivariate analysis due to unreliable documentation in the control cohort. P-values < 0.05 were considered statistically significant.

## Results

### RT is associated with an increased prevalence of CHIP

The study cohort included 736 patients with a cancer diagnosis and history of RT. Among them, 168 patients (22.8%) had a mutation in at least one CHIP gene. The median targeted sequencing coverage depth was 2,078x (range 519 - 4,106). The incidence of CHIP in post-RT patients increased with age, similar to previously reported CHIP prevalence in larger population studies^9^. However, compared to the control cohort of patients without exposure to radiation, the probability of harboring a CHIP mutation is increased in RT patients (Figure 1A). For example, at age 70, the probability of CHIP in the RT cohort vs. the control cohort is 32.4% vs. 30.5%, respectively. After adjusting for the patient’s age, sex, race, and chemotherapy exposure, RT is an independent risk factor for developing CHIP (OR 1.39, 95% CI 1.08-1.77, p=0.009 (Figure 1B). The median interval time between RT and CHIP detection in blood sample was 3.2 years.

**Figure 1:**
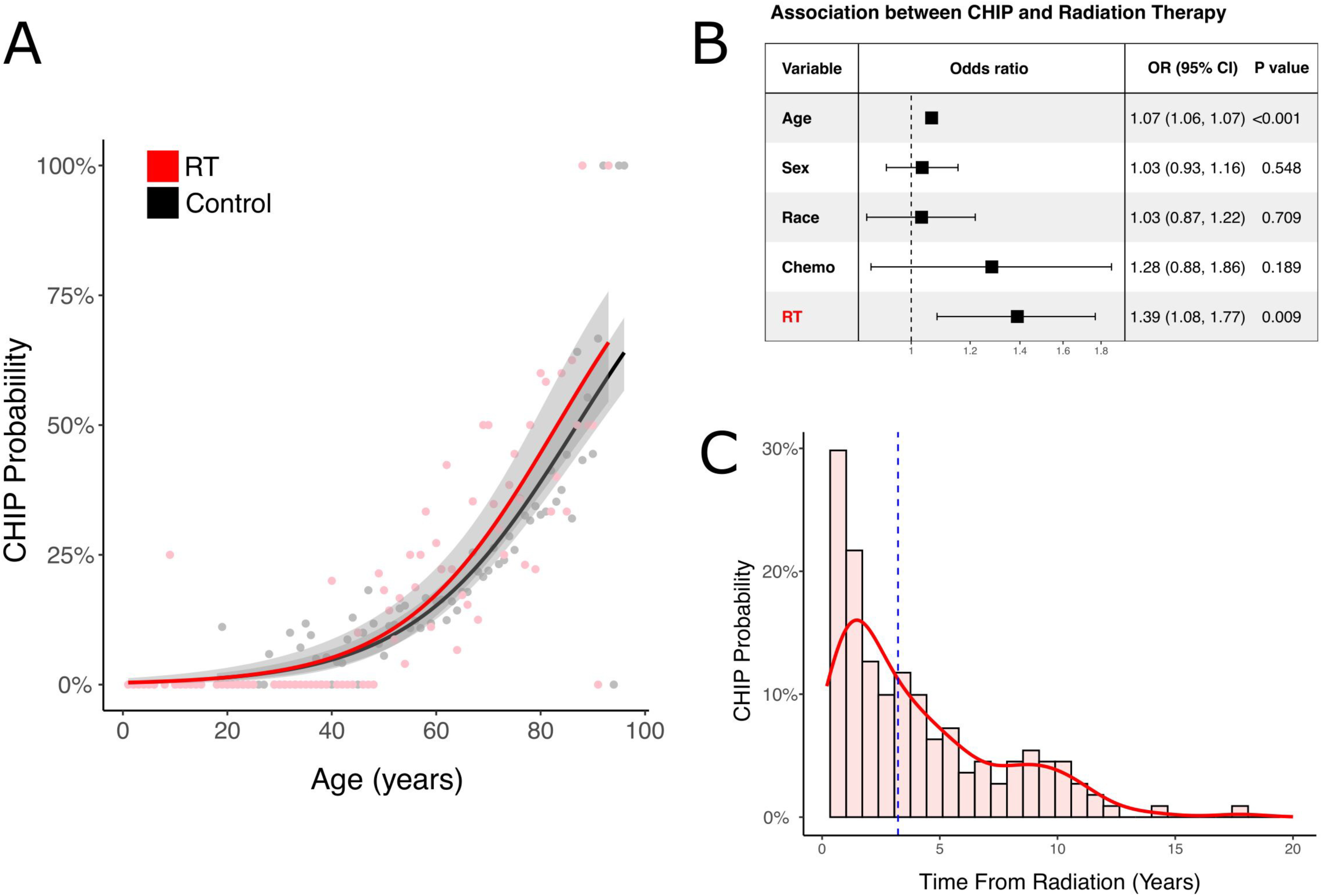
RT is associated increased prevalence of CHIP. **A.** Prevalence of CHIP based on age. Shaded region represents 95% confidence interval. Each dot indicates the probability of CHIP at specific age. **B.** Forest plot of the risk of CHIP among combined cohort of patients with and without exposure of radiation therapy. **C.** CHIP probability based on time interval between radiation therapy and blood sample collection. Red bars represent histograms of CHIP frequency based on time from radiation. Red line represents estimated probability based on the distribution of frequency.

### Specific CHIP Driver Genes are Enriched after RT

The most frequently mutated CHIP genes were *DNMT3A* (13.6%), *TET2* (5.7%), and *PPM1D* (4.1%) (Figure 2A). Compared to the control cohort of patients without a history of radiation exposure, the proportion of *DNMT3A*, *PPM1D*, *TP53*, and *BRCC3* mutations were significantly increased (13.6% vs 11.0%, p<0.05 for *DNMT3A*, 4.1% vs. 1.7%, p<0.001 for *PPM1D*, 2.4% vs. 0.9%, p<0.001 for *TP53*, and 0.7% vs. 0.2% for *BRCC3*, p<0.01). Comparison of other CHIP genes did not show a significant difference. There was no significant difference in VAF distribution between RT patients and the control cohort (Figure 2B). The types of mutations varied among individual CHIP genes, including nonsynonymous single nucleotide variants (SNV), frameshift insertions or deletions, splicing mutations, and stop-gain mutations (Figure 2C). This contradicted prior studies which identified frequent short deletions as a post-radiation mutational signature.^31,32^ Compared to the control cohort, we found no significant difference in the number of short frameshift insertions and deletions in RT patients when comparing the RT cohort and control cohort (Figure 2D).

**Figure 2:**
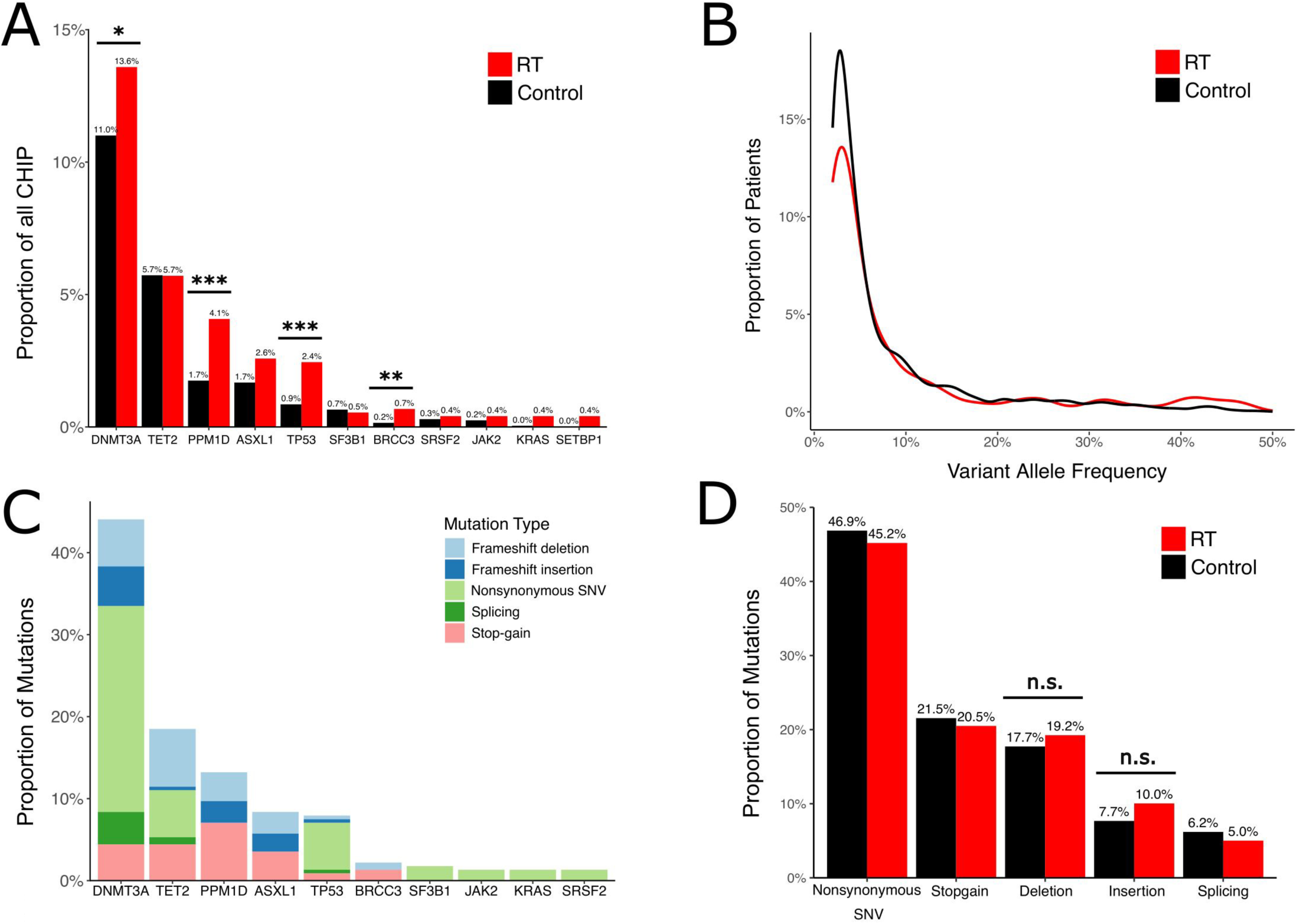
Specific CHIP Driver Genes are Enriched after RT. **A.** Prevalence of CHIP mutations in patients after RT compared to a control cohort without exposure to RT. *Chi-square test, p<0.05, ***chi-square test, p<0.001. **B**. Variant allele frequency detected in patients after RT compared to a control cohort without exposure to RT. **C.** Mutation types of individual CHIP genes among patients after radiation therapy. Only 10 CHIP genes with most mutations are shown. SNV, single-nucleotide variants. **D.** Comparison of type of mutations between the radiation cohort and the control cohort. Chi-square test, *p<0.05, **p<0.01, ***p<0.001

### CHIP risks based on cancer types

The RT cohort captured a total of 29 cancer types, with the most common being breast (n=144, 19.6%), head and neck (n=142, 19.3%), prostate (n=92, 12.5%), lung (n=88, 12.0%), and brain (n=67, 9.1%) cancers (Figure 3A). There was enrichment of specific CHIP mutations among each cancer type. For example, *PPM1D* mutations were enriched in lung, head and neck, and breast cancer, but not other cancer types (Figure 3B). Among included cancer types, lung cancer patients had the highest prevalence of CHIP (40%), followed by prostate cancer (36%) and skin cancer (24%) (Figure 3C). After adjusting for age, sex, race, smoking history, and chemotherapy, lung cancer had a significantly higher odds ratio of CHIP (OR 2.10, 95% CI 1.23-3.59, p=0.006) (Figure 3D).

**Figure 3:**
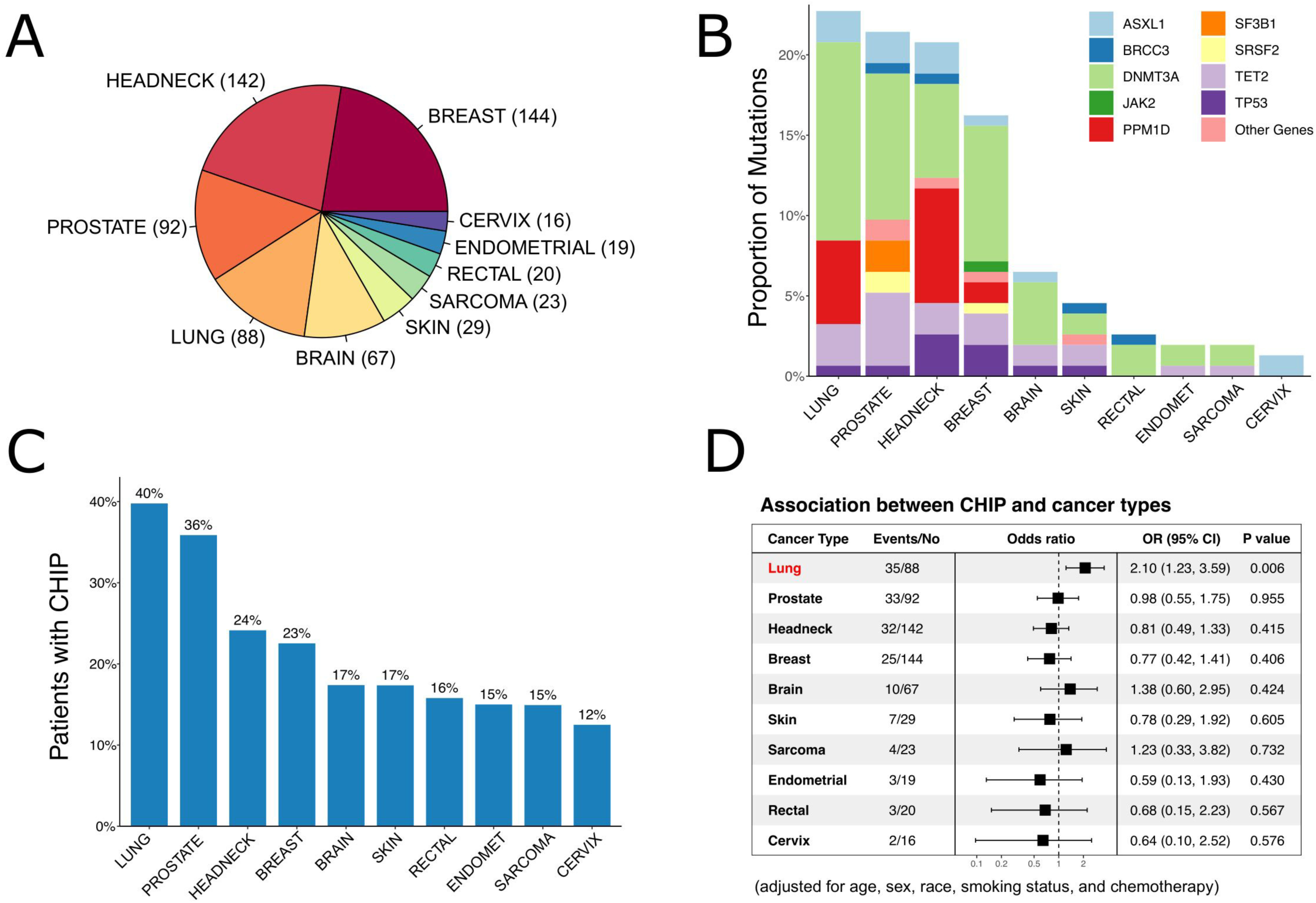
CHIP Risks Differs Among Cancer Types. **A.** Distribution of cancer types in the RT cohort. Numbers in parentheses represent number of patients included in each cancer type. **B.** Proportion of individual CHIP genes in each cancer type. Only 10 most common cancer types are shown. **C.** Proportion of patients with CHIP in each cancer type. **D.** Forest plot of association between CHIP risks and each cancer type. Odds ratios are adjusted for co-variates including age, sex, race, smoking status, and chemotherapy.

### CHIP risks and radiation parameters

We identified a positive correlation between the risk of CHIP and the dose of radiation received. This risk of CHIP is most significantly correlated with biological equivalent dose, which quantifies the expected biologic effect after adjusting for different radiation doses and fractionations, compared to radiation dose without adjustment or equivalent dose in 2Gy per fraction (Figure 4A). We investigated CHIP risks after treatment with different radiation techniques (e.g., conventional vs. stereotactic RT) and radiation modalities (e.g., photon vs. electron RT). There was a significant association between stereotactic radiation and CHIP prevalence (OR 2.24, 95% CI 1.14-4.33, p=0.02) (Figure 4B). We also examined the relationship between the irradiated body site and the risk of developing CHIP. Most patients received radiation to the body site of the primary tumor (Figure 4C). We did not find a significant difference in CHIP prevalence among different body sites irradiated (Figure 4D).

**Figure 4:**
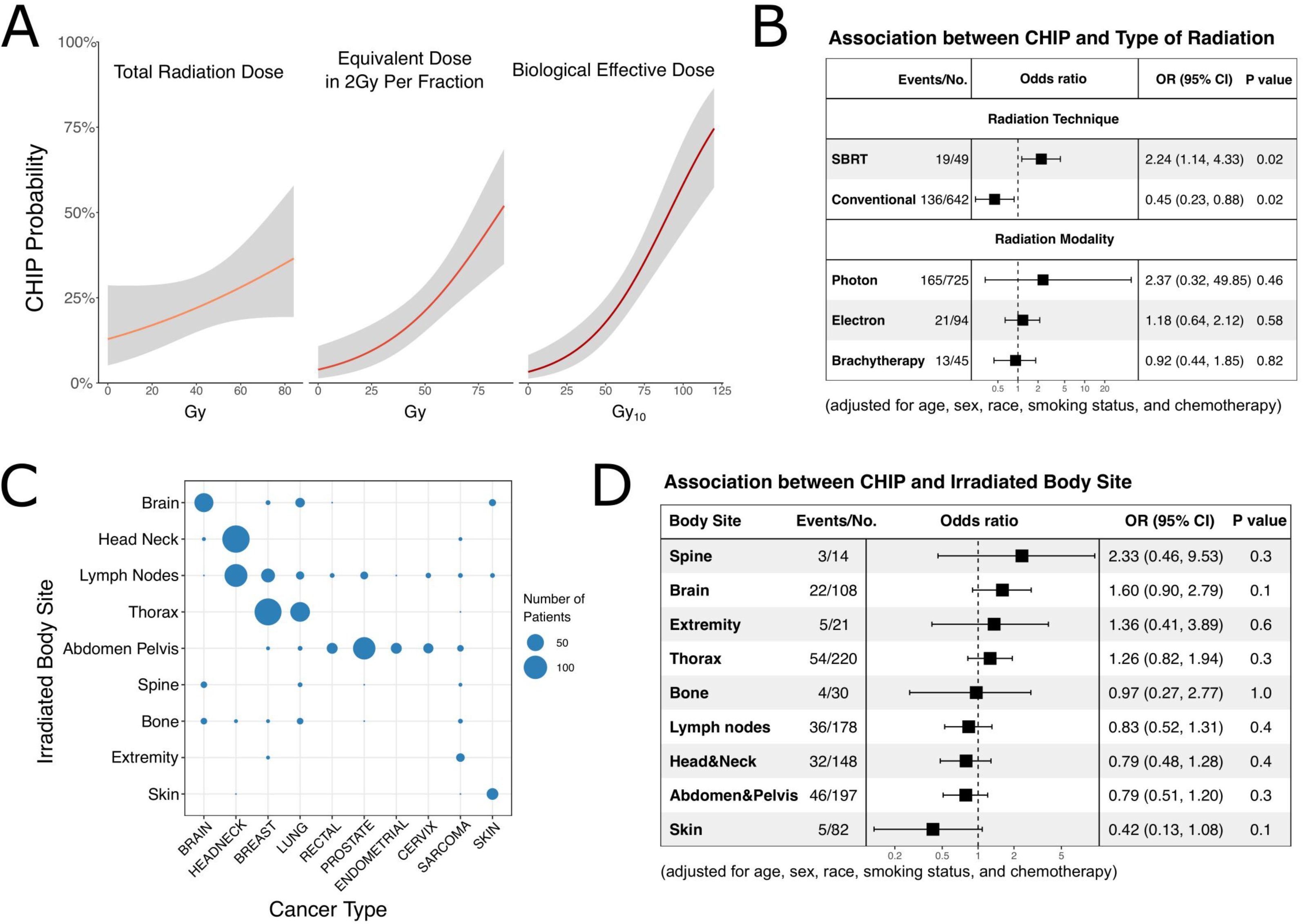
CHIP Risks Correlates with Radiation Dose and Stereotactic Technique. **A.** Relationship between CHIP probability and total radiation dose (left panel), equivalent dose in 2Gy per fraction (middle panel), and biological effective dose (right panel). Biological effective dose quantifies the expected biologic effect of different radiation dose fractionation. **B.** Forest plot of association between CHIP risks and radiation technique and modality. Odds ratios are adjusted for co-variates including age, sex, race, smoking status, and chemotherapy. **C.** Correlation between irradiated body site and cancer type. Size of each dot represents number of patients. **D.** Forest plot of association between CHIP risks and irradiated body sites. Odds ratios are adjusted for co-variates including age, sex, race, smoking status, and chemotherapy.

## Discussion

Previous studies of cancer therapy have alluded to RT as a risk factor for developing CHIP, though the exact magnitude by which RT influences the risk of developing CHIP and relevant radiation parameters that affect such risk are not well described. This is partially due to the challenges associated with collecting detailed RT data, such as radiation dose, treatment sites, and radiation techniques. Here we captured the landscape of RT-related CHIP in 736 cancer patients within a rich genotype-phenotype ecosystem based on a large institutional DNA biobank linked to the EHR that contains patient-level RT data. We found that the incidence of CHIP in post-RT patients is significantly increased compared to patients without radiation exposure in an age-independent manner. We showed that RT-related CHIP is enriched for specific CHIP driver genes. We also found that the risk of developing RT-related CHIP is the highest in primary lung cancer patients and correlates with radiation dose and stereotactic radiation technique.

Some of our findings are consistent with previous studies investigating cancer therapy-related CHIP. This includes a predilection for mutations in the DNA damage repair genes such as *TP53*, *PPM1D*, and *CHEK2*.^2,4,24,33^ It is not entirely clear why cancer therapy preferentially predisposes patients to clonal populations harboring these mutations. One hypothesis is that mutations in these genes confer resistance to cytotoxic DNA damaging therapy and improve hematopoietic stem cell survival, even without specific effect on stem cell renewal.^34^ This hypothesis is supported by evidence in cell lines containing gain-of-function *PPM1D* mutations outcompeting normal cells in the presence of cisplatin.^35^ This survival advantage was predominantly mediated by increased resistance to cisplatin-induced apoptosis. Although similar experiments have not been performed with radiation instead of cisplatin, diffuse intrinsic pontine glioma with *PPM1D* mutation has been shown to resist DNA damaging effects of ionizing radiation.^36^ This suggests that HSC clones containing *PPM1D* mutations may share similar mechanisms of survival advantage. CHIP associated with DNA damage repair mutations carries significant clinical implications. Among CHIP driver genes, *TP53* carries one of the highest risks for subsequent acute myeloid leukemia.^37^ The leukemogenic risk from *TP53* is unique to cancer therapy, as studies suggest that oncogenic mutations in *TP53* confer little or no competitive advantage in the absence of selective pressures. ^38,39^ Mutations in *TP53* are also known to promote resistance to various cancer therapies.^40–42^ The increased prevalence of *TP53* mutations in RT patients may lead to reduced effectiveness of subsequent treatments, potentially impacting long-term outcomes.

Interestingly, stereotactic RT was found to have a significant association with CHIP prevalence compared to conventional radiation techniques. This higher risk could be attributed to the concentrated, high-dose nature of stereotactic radiation, which may induce more DNA damage in hematopoietic stem cells. A surprising finding of the study was that the irradiated body site did not affect the risk of developing CHIP. Given the primary residence of HSCs in the bone marrow space, our hypothesis was that bone irradiation, particularly spine irradiation, would carry a higher risk of developing CHIP. This rejected hypothesis suggests that the systemic effects of radiation may be more important in CHIP development than the specific location of radiation exposure and/or chemotherapy given to these patients. Further investigations using 3D dosimetric data will be needed to assess the exact volume of bone marrow that was subjected to irradiation in our cohort of patients.

The strength of our study lies in the targeted sequencing assay with deep coverage and error correction, coupled with rich clinical data. This approach enabled the detection of rare CHIP clones and association studies with detailed radiation factors. However, we recognize several limitations of our study. First, a subset of patients (13%) had received chemotherapy, which can be a contributing factor to the CHIP risk. In addition, we have only collected data on platinum agents and topoisomerase inhibitors based on previous studies showing strong association of these two cytotoxic agents with CHIP risks, but not other cytotoxic agents or other systemic agents, such as immunotherapy or targeted therapy, which could also be implicated in CHIP development. Furthermore, by the design of the biobank, we do not have longitudinal samples to test clonal expansion rate in patients with CHIP. Therefore, it is unclear whether RT-associated CHIP show similar or different clonal expansion rate compared to aging-related CHIP.

Our findings have potential implications for cancer care. Clinicians may need to consider the potential long-term effects of RT, including CHIP, during consultation and radiation planning, particularly for those patients who will receive a high biological equivalent dose. Our results also emphasize the importance of monitoring CHIP in RT patients. Our institution has established a prospective registry and biorepository to identify and monitor individuals at risk for developing CHIP.^43^ The increased mutations in *PPM1D* and *TP53* could serve as potential biomarkers for assessing the impact of RT on an individual patient’s genome. There are multiple questions that deserve attention during future research. These questions include the underlying mechanism by which RT promotes CHIP, the clonal expansion rate of the RT-related CHIP clones, the potential long-term morbidity of RT-associated CHIP in cancer patients, including impact on the cardiovascular and other organ systems, and the mortality associated with therapy-related CHIP. Answering these questions will help with the development of targeted therapies to address or prevent these somatic alterations, potentially improving long-term outcomes for cancer patients.

## Conclusion

In conclusion, this study reveals that radiation therapy is associated with a significantly increased risk of developing CHIP in cancer patients. The risk correlates with radiation dose and stereotactic techniques, with unique mutational signatures observed. These findings underscore the importance of monitoring CHIP in radiation-treated patients and warrant further investigation into long-term health implications.

## Data Availability

All data produced in the present study are available upon reasonable request to the authors.

## Acknowledgements

This study received funding from the Vanderbilt Institute for Clinical and Translational Research Grant, American Cancer Society IRG Pilot Grant, and Vanderbilt Breast SPORE CEP Grant; NIH grant 5K12CA090625 (L.Y.L.); NIH grant DP5 OD029586, a Burroughs Wellcome Fund Career Award for Medical Scientists, an E.P. Evans Foundation grant, a RUNX1 Research Program grant, a Pew Charitable Trusts and Alexander and Margaret Stewart Trust Pew-Stewart Scholar for Cancer Research award, a Vanderbilt University Medical Center Brock Family Endowment grant, and a Young Ambassador Award (A.G.B.).

## Authorship Contributions

Study conceptualization: S.A.C., A.G.B., L.Y.L, E.T.S., and B.H.P.

Data collection: S.A.C., S.S.O., and C.A.B.

Data analysis: T.M.M., Y.P., Y.L., and C.A.B.

Manuscript preparation: S.A.C., LY.L., C.V., A.K., S.C.R., S.C., T.J.O., D.M.R., A.G.B., and E.T.S.

## Supplemental Figure Legends

**Supplemental Figure 1. A.** Number of CHIP mutations present in each patient. Patients without any CHIP mutations are not shown. **B**. Distribution of types of single nucleotide base-pair changes seen among RT patients with CHIP.

**Supplemental Figure 2. A.** Distribution of cancer stage within each cancer type. Only 10 most common cancer types are shown. **B.** Proportion of patients in the RT cohort that received chemotherapy (limited to platinum agents and topoisomerase inhibitors).

## References

1. Lustberg MB, Kuderer NM, Desai A, Bergerot C, Lyman GH. Mitigating long-term and delayed adverse events associated with cancer treatment: implications for survivorship. Nat Rev Clin Oncol. 2023;20(8):527–542. doi:10.1038/s41571-023-00776-9

2. Bolton KL, Ptashkin RN, Gao T, et al. Cancer therapy shapes the fitness landscape of clonal hematopoiesis. Nat Genet. 2020;52(11):1219–1226. doi:10.1038/s41588-020-00710-0

3. Boucai L, Falcone J, Ukena J, et al. Radioactive Iodine-Related Clonal Hematopoiesis in Thyroid Cancer Is Common and Associated With Decreased Survival. J Clin Endocrinol Metab. 2018;103(11):4216–4223. doi:10.1210/jc.2018-00803

4. Coombs CC, Zehir A, Devlin SM, et al. Therapy-Related Clonal Hematopoiesis in Patients with Non-hematologic Cancers Is Common and Associated with Adverse Clinical Outcomes. Cell Stem Cell. 2017;21(3):374–382.e4. doi:10.1016/j.stem.2017.07.010

5. Dawoud AAZ, Tapper WJ, Cross NCP. Clonal myelopoiesis in the UK Biobank cohort: ASXL1 mutations are strongly associated with smoking. Leukemia. 2020;34(10):2660–2672. doi:10.1038/s41375-020-0896-8

6. Mencia-Trinchant N, MacKay MJ, Chin C, et al. Clonal Hematopoiesis Before, During, and After Human Spaceflight. Cell Rep. 2020;33(10):108458. doi:10.1016/j.celrep.2020.108458

7. Steensma DP, Bejar R, Jaiswal S, et al. Clonal hematopoiesis of indeterminate potential and its distinction from myelodysplastic syndromes. Blood. 2015;126(1):9–16. doi:10.1182/blood-2015-03-631747

8. Jaiswal S, Fontanillas P, Flannick J, et al. Age-related clonal hematopoiesis associated with adverse outcomes. New England Journal of Medicine. 2014;371(26):2488–2498. doi:10.1056/NEJMoa1408617

9. Bick AG, Weinstock JS, Nandakumar SK, et al. Inherited causes of clonal haematopoiesis in 97,691 whole genomes. Nature. 2020;586(7831):763-768. doi:10.1038/s41586-020-2819-2

10. Jaiswal S, Ebert BL. Clonal hematopoiesis in human aging and disease. Science. 2019;366(6465). doi:10.1126/science.aan4673

11. Corces-Zimmerman MR, Majeti R. Pre-leukemic evolution of hematopoietic stem cells: the importance of early mutations in leukemogenesis. Leukemia. 2014;28(12):2276–2282. doi:10.1038/leu.2014.211

12. Abelson S, Collord G, Ng SWK, et al. Prediction of acute myeloid leukaemia risk in healthy individuals. Nature. 2018;559(7714):400–404. doi:10.1038/s41586-018-0317-6

13. Young AL, Tong RS, Birmann BM, Druley TE. Clonal hematopoiesis and risk of acute myeloid leukemia. Haematologica. 2019;104(12):2410–2417. doi:10.3324/haematol.2018.215269

14. Jaiswal S, Natarajan P, Silver AJ, et al. Clonal Hematopoiesis and risk of atherosclerotic cardiovascular disease. New England Journal of Medicine. 2017;377(2):111–121. doi:10.1056/NEJMoa1701719

15. Fuster JJ, MacLauchlan S, Zuriaga MA, et al. Clonal hematopoiesis associated with TET2 deficiency accelerates atherosclerosis development in mice. Science (1979). 2017;355(6327):842–847. doi:10.1126/science.aag1381

16. Dorsheimer L, Assmus B, Rasper T, et al. Association of Mutations Contributing to Clonal Hematopoiesis with Prognosis in Chronic Ischemic Heart Failure. JAMA Cardiol. 2019;4(1):25–33. doi:10.1001/jamacardio.2018.3965

17. Bhattacharya R, Zekavat SM, Haessler J, et al. Clonal Hematopoiesis Is Associated With Higher Risk of Stroke. Stroke. 2022;53(3):788–797. doi:10.1161/STROKEAHA.121.037388

18. Miller PG, Qiao D, Rojas-Quintero J, et al. Association of clonal hematopoiesis with chronic obstructive pulmonary disease. Blood. 2022;139(3):357–368. doi:10.1182/blood.2021013531

19. Bolton KL, Koh Y, Foote MB, et al. Clonal hematopoiesis is associated with risk of severe Covid-19. Nat Commun. 2021;12(1):5975. doi:10.1038/s41467-021-26138-6

20. Vlasschaert C, Robinson-Cohen C, Chen J, et al. Clonal hematopoiesis of indeterminate potential is associated with acute kidney injury. Nat Med. 2024;30(3):810–817. doi:10.1038/s41591-024-02854-6

21. Wong WJ, Emdin C, Bick AG, et al. Clonal haematopoiesis and risk of chronic liver disease. Nature. 2023;616(7958):747–754. doi:10.1038/s41586-023-05857-4

22. Agrawal M, Niroula A, Cunin P, et al. TET2-mutant clonal hematopoiesis and risk of gout. Blood. 2022;140(10):1094–1103. doi:10.1182/blood.2022015384

23. Fuster JJ, Zuriaga MA, Zorita V, et al. TET2-Loss-of-Function-Driven Clonal Hematopoiesis Exacerbates Experimental Insulin Resistance in Aging and Obesity. Cell Rep. 2020;33(4). doi:10.1016/j.celrep.2020.108326

24. Singh A, Mencia-Trinchant N, Griffiths EA, et al. Mutant PPM1D- and TP53-Driven Hematopoiesis Populates the Hematopoietic Compartment in Response to Peptide Receptor Radionuclide Therapy. JCO Precis Oncol. 2022;6:e2100309. doi:10.1200/PO.21.00309

25. Anthony KM, Collins JM, Love SAM, et al. Radon Exposure, Clonal Hematopoiesis, and Stroke Susceptibility in the Women’s Health Initiative. Neurology. 2024;102(2):e208055. doi:10.1212/WNL.0000000000208055

26. Roden D, Pulley J, Basford M, et al. Development of a Large-Scale De-Identified DNA Biobank to Enable Personalized Medicine. Clin Pharmacol Ther. 2008;84(3):362–369. doi:10.1038/clpt.2008.89

27. Pulley J, Clayton E, Bernard GR, Roden DM, Masys DR. Principles of human subjects protections applied in an opt-out, de-identified biobank. Clin Transl Sci. 2010;3(1):42–48. doi:10.1111/j.1752-8062.2010.00175.x

28. Mack T, Vlasschaert C, von Beck K, et al. Cost-effective and scalable clonal hematopoiesis assay provides insight into clonal dynamics. medRxiv. Published online November 9, 2023. doi:10.1101/2023.11.08.23298270

29. Mack T, Vlasschaert C, von Beck K, et al. Cost-Effective and Scalable Clonal Hematopoiesis Assay Provides Insight into Clonal Dynamics. J Mol Diagn. Published online April 6, 2024. doi:10.1016/j.jmoldx.2024.03.007

30. Vlasschaert C, Mack T, Heimlich JB, et al. A practical approach to curate clonal hematopoiesis of indeterminate potential in human genetic data sets. Blood. 2023;141(18):2214–2223. doi:10.1182/blood.2022018825

31. Behjati S, Gundem G, Wedge DC, et al. Mutational signatures of ionizing radiation in second malignancies. Nat Commun. 2016;7:12605. doi:10.1038/ncomms12605

32. Kocakavuk E, Anderson KJ, Varn FS, et al. Radiotherapy is associated with a deletion signature that contributes to poor outcomes in patients with cancer. Nat Genet. 2021;53(7):1088–1096. doi:10.1038/s41588-021-00874-3

33. Reed SC, Croessmann S, Park BH. CHIP Happens: Clonal Hematopoiesis of Indeterminate Potential and Its Relationship to Solid Tumors. Clin Cancer Res. 2023;29(8):1403–1411. doi:10.1158/1078-0432.CCR-22-2598

34. Florez MA, Tran BT, Wathan TK, DeGregori J, Pietras EM, King KY. Clonal hematopoiesis: Mutation-specific adaptation to environmental change. Cell Stem Cell. 2022;29(6):882–904. doi:10.1016/j.stem.2022.05.006

35. Hsu JI, Dayaram T, Tovy A, et al. PPM1D Mutations Drive Clonal Hematopoiesis in Response to Cytotoxic Chemotherapy. Cell Stem Cell. 2018;23(5):700–713.e6. doi:10.1016/j.stem.2018.10.004

36. Akamandisa MP, Nie K, Nahta R, Hambardzumyan D, Castellino RC. Inhibition of mutant PPM1D enhances DNA damage response and growth suppressive effects of ionizing radiation in diffuse intrinsic pontine glioma. Neuro Oncol. 2019;21(6):786–799. doi:10.1093/neuonc/noz053

37. Abelson S, Collord G, Ng SWK, et al. Prediction of acute myeloid leukaemia risk in healthy individuals. Nature. 2018;559(7714):400–404. doi:10.1038/s41586-018-0317-6

38. McKerrell T, Vassiliou GS. Aging as a driver of leukemogenesis. Sci Transl Med. 2015;7(306):306fs38. doi:10.1126/scitranslmed.aac4428

39. Wong TN, Ramsingh G, Young AL, et al. Role of TP53 mutations in the origin and evolution of therapy-related acute myeloid leukaemia. Nature. 2015;518(7540):552–555. doi:10.1038/nature13968

40. Alam SK, Yadav VK, Bajaj S, et al. DNA damage-induced ephrin-B2 reverse signaling promotes chemoresistance and drives EMT in colorectal carcinoma harboring mutant p53. Cell Death Differ. 2016;23(4):707–722. doi:10.1038/cdd.2015.133

41. Sampath J, Sun D, Kidd VJ, et al. Mutant p53 cooperates with ETS and selectively up-regulates human MDR1 not MRP1. J Biol Chem. 2001;276(42):39359–39367. doi:10.1074/jbc.M103429200

42. Huang Y, Liu N, Liu J, et al. Mutant p53 drives cancer chemotherapy resistance due to loss of function on activating transcription of PUMA. Cell Cycle. 2019;18(24):3442–3455. doi:10.1080/15384101.2019.1688951

43. Shannon ML, Heimlich JB, Olson S, et al. Clonal hematopoiesis and inflammation in the vasculature: CHIVE, a prospective, longitudinal clonal hematopoiesis cohort and biorepository. Blood Adv. 2024;8(13):3453–3463. doi:10.1182/bloodadvances.2023011510

