## Supplemental Figures and Table for "Risk of Clonal Hematopoiesis of Indeterminate Potential after Cancer Radiation Therapy"

A

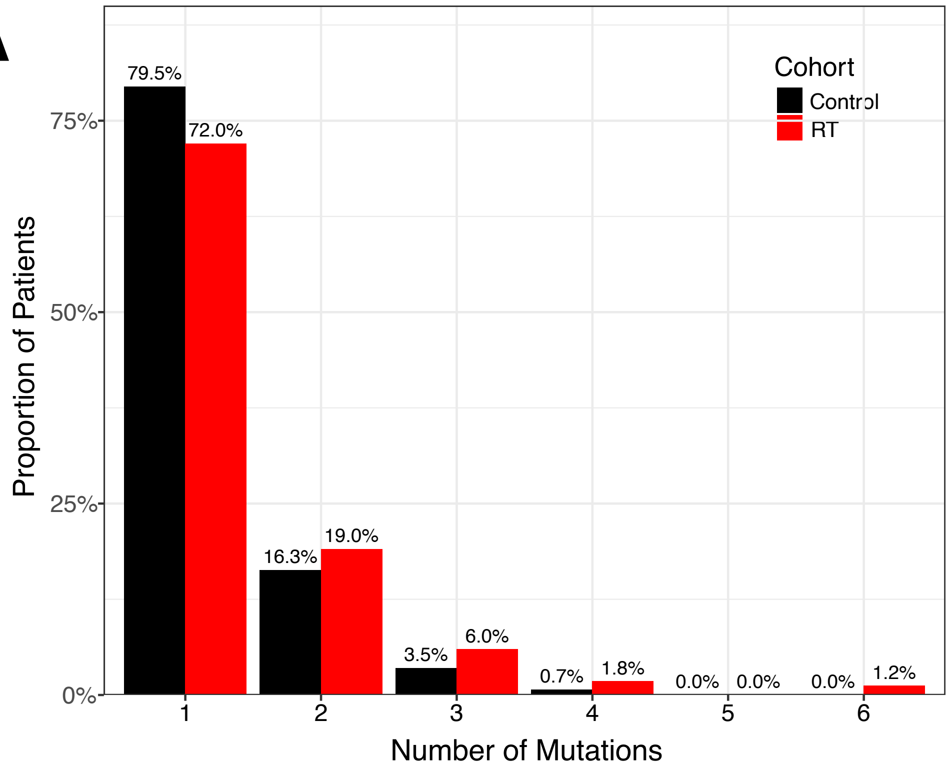
Cohort
Control
RT
79.5%
72.0%
16.3%
19.0%
3.5%
6.0%
0.7%
1.8%
0.0%
0.0%
0.0%
1.2%
75%
50%
25%
0%
Proportion of Patients
1
2
3
4
5
6
Number of Mutations

B

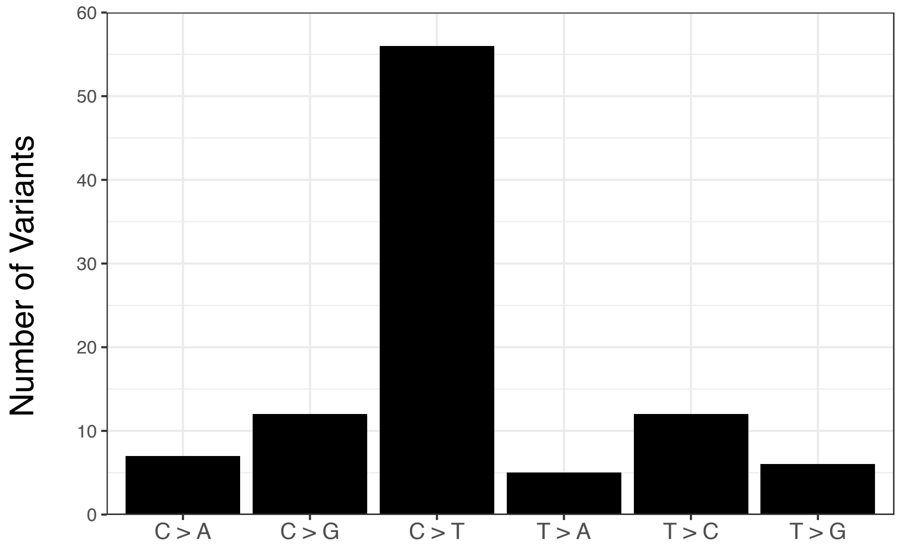
60
50
40
30
20
10
0
Number of Variants
C > A
C > G
C > T
T > A
T > C
T > G

S1

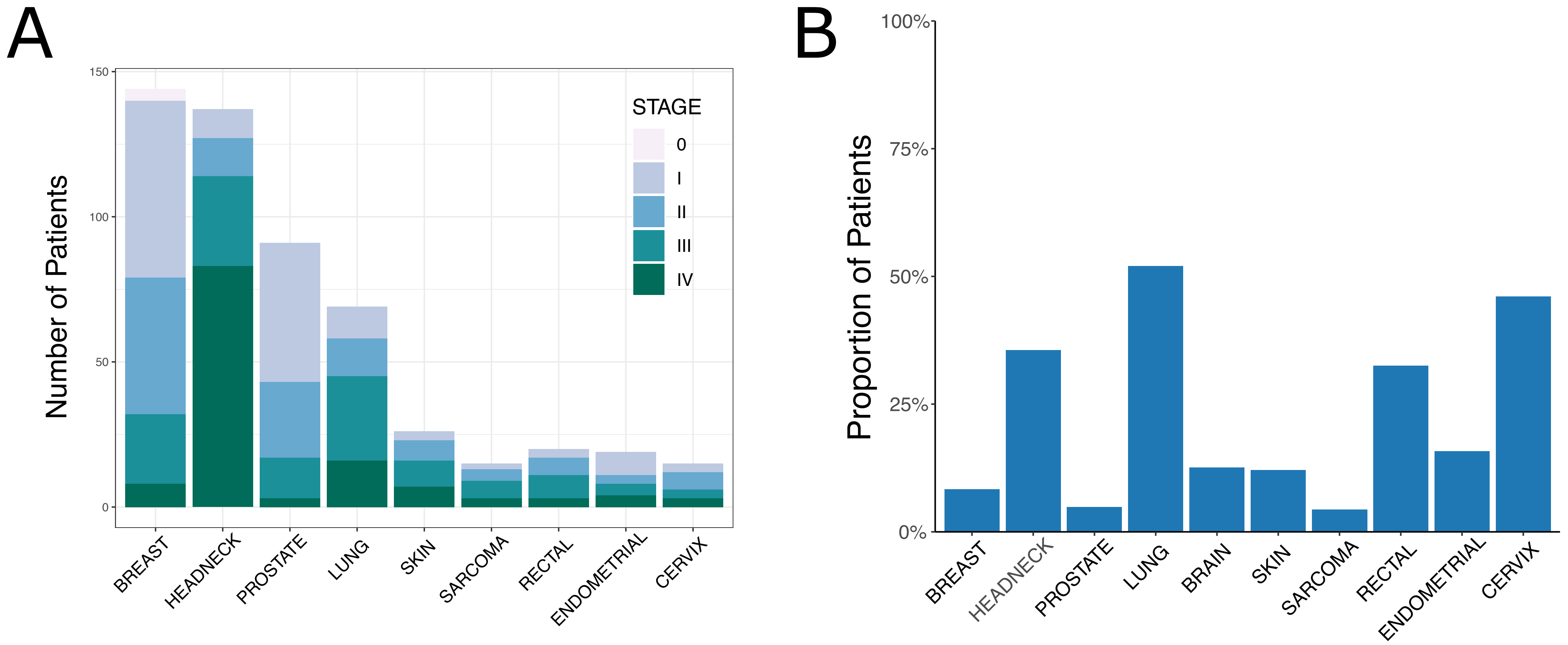
A
150
100
50
0
Number of Patients
STAGE
0
I
II
III
IV
BREAST
HEADNECK
PROSTATE
LUNG
SKIN
SARCOMA
RECTAL
ENDOMETRIAL
CERVIX
B
100%
75%
50%
25%
0%
Proportion of Patients
BREAST
HEADNECK
PROSTATE
LUNG
BRAIN
SKIN
SARCOMA
RECTAL
ENDOMETRIAL
CERVIX

S2

**Supplemental Table 1.** Current Procedural Terminology (CPT) Codes Used to Identify Radiation-Exposed Patients

| CPT Code | Associated Procedure |
|---|---|
| 77295 | 3-dimensional radiotherapy plan |
| 77301 | IMRT dose planning |
| 77306 | Teletherapy isodose plan |
| 77307 | Teletherapy isodose plan |
| 77316 | Brachytherapy isodose plan |
| 77317 | Brachytherapy isodose plan |
| 77318 | Brachytherapy isodose plan |
| 77321 | Special teletherapy port plan |
| 77371 | SRS treatment delivery |
| 77372 | SRS treatment delivery |
| 77373 | SBRT treatment delivery |
| 77385 | IMRT treatment delivery |
| 77386 | IMRT treatment delivery |
| 77401 | Radiation treatment delivery, superficial and/or orthovoltage |
| 77402 | Radiation treatment delivery |
| 77407 | Radiation treatment delivery |
| 77412 | Radiation treatment delivery |
| 77761 | Intracavitary radiation |
| 77762 | Intracavitary radiation |
| 77763 | Intracavitary radiation |
| 77767 | Under Clinical Brachytherapy Radiation Treatment |
| 77768 | Under Clinical Brachytherapy Radiation Treatment |
| 77770 | HDR Brachytherapy |
| 77771 | HDR Brachytherapy |
| 77772 | HDR Brachytherapy |
| 77778 | LDR Brachytherapy |
| 77789 | Surface application of radiation source |
